# No conclusive evidence for an association between differential observational threat learning and physical activity in everyday life

**DOI:** 10.1101/2024.05.06.24305204

**Authors:** Jannis Petalas, Madeleine Müller, Jan Haaker

## Abstract

**Background:** Exercise is well known to generally improve health status in humans and seems to be beneficial not only for physical, but also learning processes. The evidence for the impact of general physical activity on emotional learning, is however scarce. Here, we test the pre-registered hypothesis that the individual physical activity level of the past seven days is positively associated with observational threat learning, indicated by the differentiation between threat and safety cues.

**Methods:** We conducted a two day online study. 90 healthy participants (mean age = 27.82 years) engaged by completing questionnaires (State-Trait Anxiety Inventory (STAI-S/STAI-T and International Physical Activity Questionnaire (IPAQ)) on day one, followed by an observational learning task. Participants were asked to rate their discomfort, fear, and physiological response towards the CS+ and CS-both before and after the learning phase using a visual analogue scale. On day two (approximately 24 hours after day one), participants completed the STAI-S again, followed by a direct generalization task. Similar to before, participants were asked to report their discomfort, fear, and physiological response both before and after the generalization task.

To quantify the level of physical activity (PA) of the past seven days a sum score of the IPAQ was calculated. The pre-registered primary endpoint was to test for apositive association between PA and the ability to discriminate the conditioned stimuli.

**Results:** Pearson’s correlation analyses revealed no significant correlations between the combined total physical activity (PA) score and differential ratings of subjective discomfort (r = 0.11, p_corr_ = 0.150), fear (r = 0.203, p_corr_ = 0.081) or physiological responses (r = 0.145, p_corr_ = 0.17) on day 1. The same analysis revealed no significant correlations on day 2, either (differential ratings of subjective discomfort, r = 0.053, p_corr_ = 0.93, fear, r = -0.068, p_corr_ = 0.99, and physiological responses, r = -0.072, p_corr_ = 0.751 on day 2). However, we also found no sufficient evidence supporting the null hypothesis (i.e. no correlation) for the association between differential learning and PA on day 1, when applying bayesian statistics. Instead, we found a covariation between the ratings of discomfort and physical activity, as well as between ratings of fear and physical activity on day 1 and 2 within a repeated measurement ANOVA. This was supported by bayesian statistics.

**Conclusion:** Our results provided no convincing evidence for a correlation between differential observational threat learning and physical activity (as measured by the IPAQ). Future studies that provide a better control for individual physical activity are warranted.

## Introduction

Exercise is well known to generally improve health status in humans. Especially in prevention of cardiovascular, immunological or metabolic diseases, exercise has positive effects. However, exercising seems to be beneficial not only physically, but also cognitively. Over the last decades findings in biomedical research provide evidence that exercise may contribute to enhanced cognitive functions, like learning and memory (Erickson et al., 2011; Cassilhas et al., 2016; Voss et al., 2019).

The current study examines the impact of general physical exercise on emotional learning, more precisely fear conditioning. Fear conditioning is a common laboratory model to investigate associative learning processes and emotional memory retention. In differential fear conditioning, participants learn that a conditioned stimulus (CS+) is predictive for an aversive unconditioned stimulus (US). A second stimulus (CS-), is not predictive for the US. By learning the differential prediction of the US by the two CSs, the participants start expressing conditioned threat responses that are higher to the CS+ as compared to the CS-. As such, discrimination of threat and safety can be examined by the discrimination between CS+ and CS-. However, threat and safety is commonly not learned by firsthand experience of aversive outcomes (e.g. such as an US). Instead, observational learning, a form of social learning, enables us to learn about threats and safety by observing the behavior of others. When applying an observational threat learning paradigm, the participants observe a demonstrator model that is exposed to the CS+, which is followed by the US. In this case, the subject learns by observing the responses of another person to predict the occurrence of the US by the presence of CS+ and exhibits conditioned threat responses, when exposed to the CS+ themselves (Olsson & Phelps, 2004; Haaker et al., 2017).

Since various studies demonstrated a possible memory strengthening effect of physical activity (PA) (Erickson et al., 2011; Cassilhas et al., 2016; Voss et al., 2019), one possibility is that PA also contributes to an enhanced emotional learning and memory, such as the ability to differentiate threat and safety (i.e., CS+ and CS-).

To date, there has been little research on the effect of PA on fear conditioning, especially in humans. Keyan and Bryant (2019) demonstrated an enhancement of fear extinction after an acute exercise intervention in healthy participants. Interestingly, 24 hours post exercise the intervened group demonstrated significantly lower retention of conditioned fear to the CS+ (but higher responses to the CS-). Another study in women diagnosed with post-traumatic stress disorder (PTSD) used aerobic exercise as an intervention during a consolidation window of 24 hours following fear extinction learning (Crombie et al., 2021). Their results demonstrated significantly reduced threat expectancy ratings following a reinstatement. In contrast, Jacquart and colleagues (2017) could not demonstrate that exercise in rats facilitates fear extinction, long-term memory, or fear relapse tests in four distinct conditioning and extinction paradigm experiments. Moreover, another experiment by Jacquart et al. (2017) in humans with diagnosed anxiety-related disorders also failed to demonstrate enhancement in symptom improvement when adding an exercise intervention to an exposure therapy. Hence, the evidence for an enhancement of emotional learning by PA is mixed.

It is further unanswered if the discrimination between threat and safety cues via observational threat learning is associated with individual levels of PA.

In the current study, we examined the pre-registered hypothesis that the individual PA levels of the past seven days (collected via the international physical activity questionnaire, IPAQ) is positively associated with the discrimination between threat and safety cues that are learned from observation.

## Methods

### Paradigm

Participants engaged in this online study by completing questionnaires (State-Trait Anxiety Inventory (STAI-S/STAI-T)(Spielberger, 1983), presented via www.soscisurvey.de) on day one, followed by the observational learning task (conducted using PsychoPy3). During this task, participants observed a demonstrator completing two blocks of threat learning with six presentations of conditioned stimuli (CS) per block (3x CS-reinforced; 2x CS+ reinforced; 1x CS+ not reinforced). This resulted in a total of 12 trials (6x CS+, 6x CS-) with a reinforcement rate of 66%. The stimulus order was predetermined for the two blocks, but the block order was randomized. An inter-trial interval (ITI) was presented after each CS, with a randomized duration between 4 and 6 seconds. Participants were asked to rate their discomfort, fear, and physiological response towards the CS+ and CS-both before and after the learning phase using a visual analogue scale (e.g., “How much discomfort do you feel when confronted with this picture [CS]? 0 (no discomfort) - 10 (much discomfort)”; similar scales were used for fear and physiological response).

On day two (approximately 24 hours after day one), participants completed the STAI-S again, followed by a direct generalization task. The results of the generalization task are not presented here, as they are part of a separate project conducted with a different sample. Prior to the generalization task, participants were instructed to wear headphones and informed that they might hear an unpleasant sound (US). They were asked to adjust their computer volume to the maximum setting, and a test sound (“beep”) was played. If they couldn’t hear the sound, they were instructed to check their headphones and volume settings and retry the test until they could hear the sound. However, no aversive sound was actually presented during the task. The generalization task comprised three blocks in which five generalization stimuli (GS) along with the two learned CSs were presented pseudorandomly. An ITI was presented after each CS and GS, with a randomized duration between 4 and 6 seconds. Before the second and third blocks, participants were shown two videos featuring the reinforced CS+ and the CS-as a reminder. Participants were asked to rate how safe/dangerous they perceived their situation when confronted with each stimulus on a scale of 1-4 (1 = safe/4 = dangerous) during each presentation of the CS/GS. Similar to before, participants were asked to report their discomfort, fear, and physiological response both before and after the generalization task.

### Stimulus material

Images depicting either yellow or blue doorbells were used as conditioned stimuli (CSs) (based on Skversky-Blocq et al., 2021). The colors of the CS+ and CS-were counterbalanced. Observed CSs were presented in videos lasting 12 seconds, featuring a male demonstrator wearing headphones while looking at a monitor displaying either the CS+ or the CS-. The onset of the observed US was indicated by the demonstrator’s facial reaction to an unpleasant sound, which occurred seven seconds into the video and lasted for one second. Generalized stimuli consisted of five doorbells with a color gradient ranging from yellow to blue (based on Skversky-Blocq et al., 2021).

### Pre-registration

The protocol and analyses were pre-registered. See https://osf.io/egnma

### Subjects

We enrolled healthy individuals aged 18 to 65 years in this online study until we achieved our predetermined sample size of over 89 participants for the observational learning task. Participation required access to a computer with headphones. Individuals were excluded from analysis if they did not complete all parts of the study (observational learning, direct generalization), or if the time gap between the observational learning (ACQ on day 1) and generalization (GEN on day 2) tasks was less than 18 hours or more than 30 hours (i.e., 24 hours ±6 hours). Moreover, participants were excluded if they consistently provided over 90% uniform ratings across all stimuli types during the safe/danger ratings on day 2 (i.e., clicking through consistently). Initially, we recruited 153 participants. After applying the exclusion criteria, our final sample size was 90 participants (66 female, 23 male, 1 diverse). Participation was remunerated with €10. Participants provided demographic information including age (mean age = 27.82 years, sd = 5.62, min=19 years, max=53 years), gender, alcohol consumption (mean alcohol consumption = 1.041 units per week, sd = 1.578, min = 0/week, max = 8/week), coffee consumption (mean coffee consumption = 1.236 units per day, sd = 1.092, min = 0/day, max = 5/day), and smoking status (79 non-smokers, 11 smokers).

### Methods main effects

Ratings for fear, discomfort and physiological response for day 1 and 2 were entered into a repeated measures ANOVA with the factor Stimulus (2 levels: CS+, CS-) and Time (2 levels: before learning, after learning).

Post-hoc tests that were indicative of observational threat learning on day 1 were defined (as pre-registered) as comparing ratings between the CS+ and the CS-, after learning, when compared to before (Formula: (CS+ - CS-) after acquisition) vs. ((CS+ - CS-) before acquisition).

Post-hoc tests for the retrieval of observational learning on day 2 were defined as comparing ratings to the CS+ vs. CS-before generalization on day 2.

Pearson’s correlation analyses were calculated for each questionnaire (anxious temperament: STAI Trait; momentary anxiety: STAI state; emotional empathy: BEES) and the differential score for observational learning (Formula: (CS+ - CS-) after acquisition) vs. ((CS+ - CS-) before acquisition) for each rating measure (fear; discomfort; physiological arousal). Each p-value was corrected for multiple comparisons (i.e. three ratings measures) using Bonferroni-Holm method.

### Data analysis PA

For statistical analysis a sum score of physical activity was calculated. Physical activity (PA) score was calculated by summing the durations of PA (in minutes) and multiplying them with the frequency (see IPAQ Guidelines, 2004). The combined total PA score is a non-weighted score, treating all forms of PA (walking, moderate-intensity and vigorous-intensity activity) equally.

Total PA time per week was truncated to a maximum of 960 minutes (16 hours), as suggested by the IPAQ Guidelines (2004). Total PA time per day was truncated (re-coded) to be equal to 240 minutes, also suggested by the IPAQ Guidelines (2004) to normalize the distribution of levels of PA.

The primary endpoint for the current research hypothesis was an association between PA and the ability to discriminate the conditioned stimuli. To determine the hypothesized association between the combined total PA score and the CS differences a pearson’s correlation was calculated. As mentioned above, CS differences consisted of the difference between ratings to the CS+ and the CS-after learning, subtracted by the difference between the CSs at baseline, before learning ((CS+ > CS-) after learning > (CS+ > CS-) before learning). Calculations were executed separately for the ratings of discomfort and fear, as well as the physiological response for day 1 and day 2.

All tests were calculated one-tailed for a positive correlation, based on the pre-registered hypothesis. The Bonferroni-Holm method was applied to correct family-wise error rates.

Additionally, we examined individual PA as a covariate in the analysis of the main factors in observational learning (stimulus and time) within a type III repeated measures analysis of variance (rmANOVA). This analysis has not been pre-registered and was therefore explorative. Repeated measures factors were determined as time (pre- and post-values) and stimulus (CS- and CS+). The combined total PA score was integrated in the analysis as a covariate. Partial η^2^ were calculated to estimate effect sizes.

Bayesian statistics were used to quantify the evidence for the Null Hypothesis of the Correlation, by calculating the BF_01_.

To further evaluate statistically significant results of the rmANOVA, a Bayesian rmANOVA was calculated. This analysis included the model terms, time’,, stimulus’, as well as, time*stimulus’ interaction as a null model, which was compared to the evidence to include combined total PA as a covariate.

## Results

### Main effects

#### Day 1

Our analysis revealed successful observational learning on day 1, reflected by a Stimulus*Time interaction within all outcomes measures (fear ratings: F(1.000)=37.490; p<0.001; η^2^_p_=0.291; discomfort: F(1.000)=36.463; p<0.001; η^2^_p_=0.291; physiological response: F(1.000)=27.245; p<0.001; η^2^_p_=0.234; see table 1 & figure 1). As expected, all outcome measures indicated increasing CS-differentiation, i.e., that the ratings were higher to the CS+, as compared to the CS-, after observational learning, when compared to the ratings before (post-hoc t-test fear ratings: t(89)=-6.123; p<0.001; discomfort: t(89)=-6.038; p<0.001; physiological response: t(89)=-5.220; p<0.001; see table 3).

**Table 1.** Displays results of rmANOVA for day 1.

| | Sum of Squares | df | Mean Square | F | p | $\eta^2_p$ |
| --- | --- | --- | --- | --- | --- | --- |
| discomfort day 1 |  |  |  |  |  |  |
| time | 2.318 | 1.000 | 2.318 | 0.460 | 0.5 | 0.005 |
| stimulus | 84.214 | 1.000 | 84.214 | 30.727 | < .001 | 0.257 |
| time * stimulus | 95.962 | 1.000 | 95.962 | 36.463 | < .001 | 0.291 |
| fear day 1 |  |  |  |  |  |  |
| time | 105.917 | 1.000 | 105.917 | 34.330 | < .001 | 0.278 |
| stimulus | 67.400 | 1.000 | 67.400 | 22.543 | < .001 | 0.202 |
| time * stimulus | 61.903 | 1.000 | 61.903 | 37.490 | < .001 | 0.291 |
| physiological response day 1 |  |  |  |  |  |  |
| time | 17.654 | 1.000 | 17.654 | 8.018 | 0.006 | 0.083 |
| stimulus | 60.801 | 1.000 | 60.801 | 24.699 | < .001 | 0.217 |
| time * stimulus | 44.254 | 1.000 | 44.254 | 27.245 | < .001 | 0.234 |
| Note. Sphericity corrections not available for factors with 2 levels. |  |  |  |  |  |  |
| Note. Type III Sum of squares |  |  |  |  |  |  |

**Table 2.**
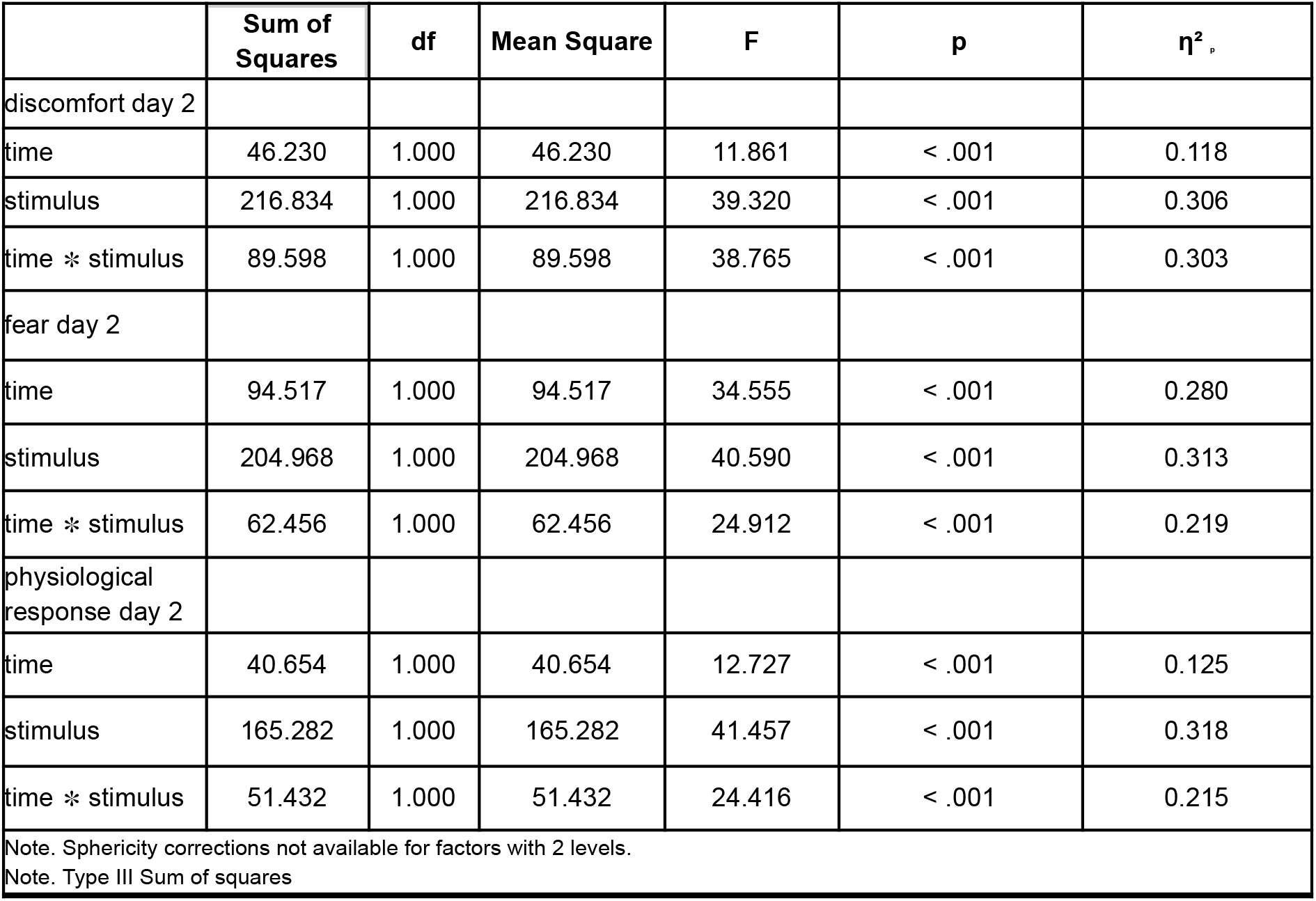
Displays the results of the rmANOVA for day 2.

**Table 3.** Displays post-hoc t-test stimulus*time for discomfort, fear and physiological response.

| Paired Samples T-Test |  |  |  |  |  |  |  |
| --- | --- | --- | --- | --- | --- | --- | --- |
|  |  |  |  |  |  | 95% CI for Cohen's d |  |
| Measure 1 | Measure 2 | t | df | p | Cohen's d | Lower | Upper |
| disc_diff_d1_pre | - disc_diff_d1_post | -6.038 | 89 | < .001 | -0.637 | -0.862 | -0.408 |
| fear_diff_d1_pre | - fear_diff_d1_post | -6.123 | 89 | < .001 | -0.645 | -0.871 | -0.417 |
| phys_diff_d1_pre | - phys_diff_d1_post | -5.220 | 89 | < .001 | -0.550 | -0.771 | -0.327 |
| disc_diff_d2_pre | - disc_diff_d2_post | -6.226 | 89 | < .001 | -0.656 | -0.883 | -0.427 |
| fear_diff_d2_pre | - fear_diff_d2_post | -4.991 | 89 | < .001 | -0.526 | -0.745 | -0.304 |
| phys_diff_d2_pre | - phys_diff_d2_post | -4.941 | 89 | < .001 | -0.521 | -0.740 | -0.299 |
| Alt.Hypothesis: Measure 1 does not equal Measure 2 |  |  |  |  |  |  |  |

**Figure 1.**
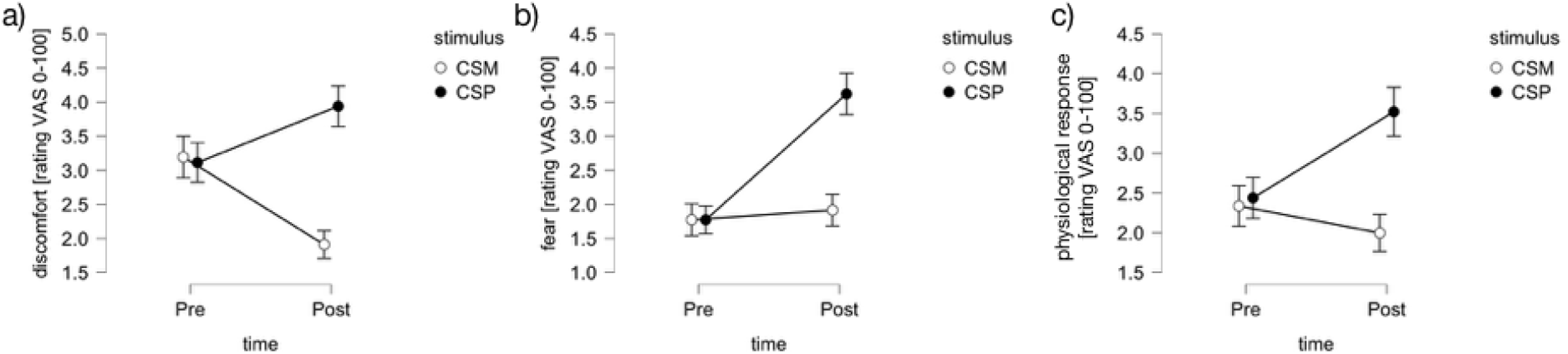
Display of the results of the rmANOVA for discomfort (a), fear (b) and physiological response (c) on day 1, pre and post observational learning. Error bars indicate standard errors of the mean.

#### Day 2

On day 2, participants retrieved the previously learned association and thus differentiated between the CS+ and the CS-within all outcome measures, indicated by a stimulus main effect (fear ratings: F(1.000)=40.590; p<0.001; η^2^_p_=0.313; discomfort: F(1.000)=39.320; p<0.001; η^2^_p_=0.306; physiological arousal: F(1.000)=41.457; p<0.001; η^2^_p_=0.318; see figure 2 & see table 2). Post-hoc tests revealed higher ratings to the CS+ as compared to the CS-for fear, discomfort and physiological arousal at the start of the experiment (post-hoc t-test retrieval fear ratings: t(89)=-3.011; p=0.003; discomfort: t(89)=-2.349; p=0.021; physiological arousal: t(89)=-3.103; p=0.003; see figure 3).

**Figure 2.**
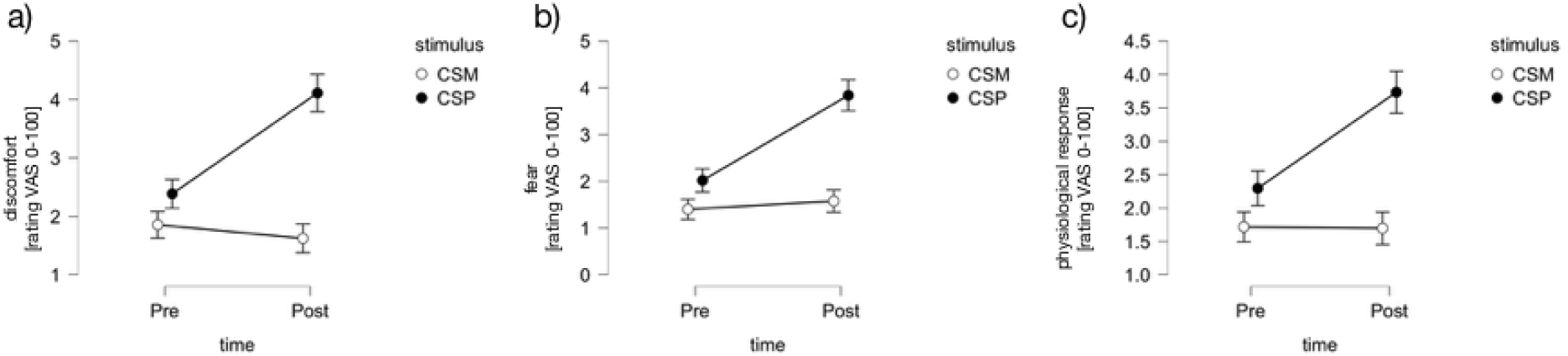
Display of the results of the rmANOVA for discomfort (a), fear (b) and physiological response (c) on day 2, pre and post observational learning. Error bars indicate standard errors of the mean.

**Figure 3.**
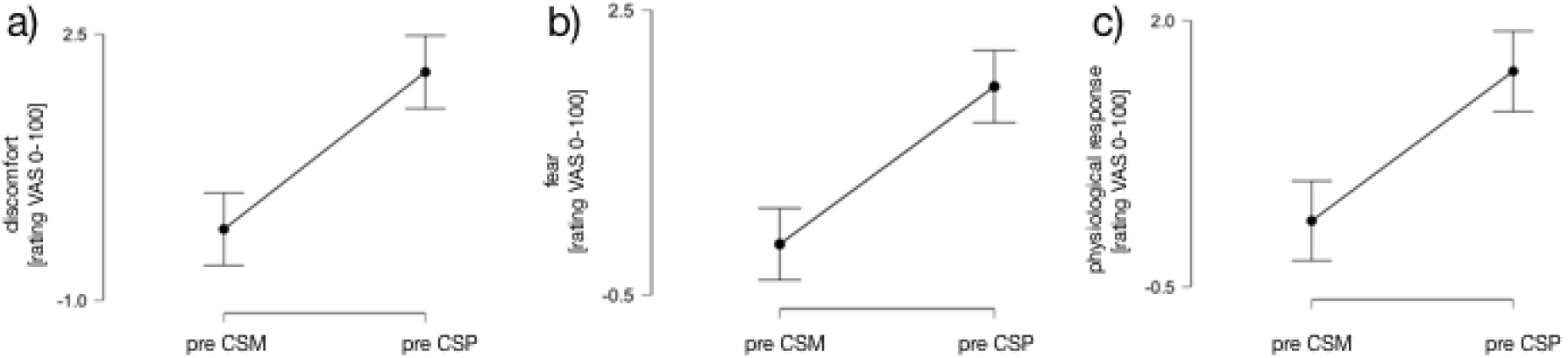
Display of the results of the post-hoc-t-test retrieval ratings for discomfort (a), fear (b) and physiological response (c) on day 2. Error bars indicate 95% confidence interval for the mean.

Additionally, the analysis indicated a stimulus*time interaction, consisting of increasing ratings to the CS+ compared to the CS-during the generalization phase (fear ratings: F(1.000)=24.912; p<0.001; η^2^_p_=0.219; discomfort: F(1.000)=38.765; p<0.001; η^2^_p_=0.303; physiological arousal: F(1.000)=24.416 p<0.001; η^2^_p_=0.215; see table 2; (post-hoc t-test stimulus*time fear ratings: t(89)=-4.991; p<0.001; discomfort: t(89)=-6.226; p<0.001; physiological arousal: t(89)=-4.941; p<0.001; see table 3).

### Correlations (Association between differential learning and anxious temperament)

Our analysis revealed a negative association between individual anxious temperament measures (STAI Trait) and differential ratings of discomfort during observational learning (((CS+ > CS-) after acquisition) vs. ((CS+ - CS-) before acquisition); r= -0.293; p_corr_=0.015; see table 4). The analysis revealed no association between differential ratings and anxious temperament (STAI Trait) on day 2 (all ps>0.663; see table 4). We further found no associations between CS differentiation and anxious state (STAI State all ps>0.081) or emotional empathy (BEES p=0.290; see table 4).

**Table 4.** Display of pearson’s correlations between differential learning, physical activity and anxious temperament measure as well as the brief emotional experience scale.

| Differential learning index ((CS+ > CS-) after acquisition/ generalization) vs. (CS+ - CS-) before acquisition/ generalization) |  | STAI level of agreement (State anxiety) | Holm-Bonferroni corrected | STAI level of agreement (Trait anxiety) | Holm-Bonferroni corrected | BEES percentage score | Holm-Bonferroni corrected |
| --- | --- | --- | --- | --- | --- | --- | --- |
| discomfort day 1 | Pearson's r | -0.185 |  | -0.293 |  | 0.077 |  |
|  | p-value | 0.081 | 0.243 | 0.005 | 0.015 | 0.471 | 0.942 |
| fear day 1 | Pearson's r | 0.051 |  | -0.010 |  | -0.013 |  |
|  | p-value | 0.631 | 0.631 | 0.925 | 0.925 | 0.900 | 0.900 |
| physiological response day 1 | Pearson's r | -0.064 |  | -0.109 |  | 0.113 |  |
|  | p-value | 0.552 | 0.999 | 0.307 | 0.921 | 0.290 | 0.870 |
| discomfort day 2 | Pearson's r | 0.010 |  | -0.016 |  | -0.045 |  |
|  | p-value | 0.926 | 0.926 | 0.881 | 0.999 | 0.671 | 2.013 |
| fear day 2 | Pearson's r | -0.046 |  | -0.047 |  | -0.028 |  |
|  | p-value | 0.666 | 0.999 | 0.663 | 0.999 | 0.797 | 0.797 |
| physiological response day 2 | Pearson's r | -0.067 |  | 0.001 |  | -0.030 |  |
|  | p-value | 0.531 | 0.999 | 0.990 | 0.990 | 0.779 | 1.558 |
| Alt. hypothesis: Correlated |  |  |  |  |  |  |  |

### Association between differential learning and physical activity

Against our hypothesis, Pearson’s correlation analyses revealed no significant correlations between the combined total physical activity (PA) score and differential ratings of subjective discomfort (r = 0.11, p_corr_ = 0.150), fear (r = 0.203, p_corr_ = 0.081) or physiological responses (r = 0.145, p_corr_ = 0.17) on day 1 (see figure 4). The same analysis revealed no significant correlations on day 2, either (differential ratings of subjective discomfort, r = 0.053, p_corr_ = 0.93, fear, r = -0.068, p_corr_ = 0.99, and physiological responses, r = -0.072, p_corr_ = 0.751 on day 2; figure 5). We further found no association between PA scores and anxious temperament, anxious state or emotional empathy (all ps > 0.066)

**Figure 4.**
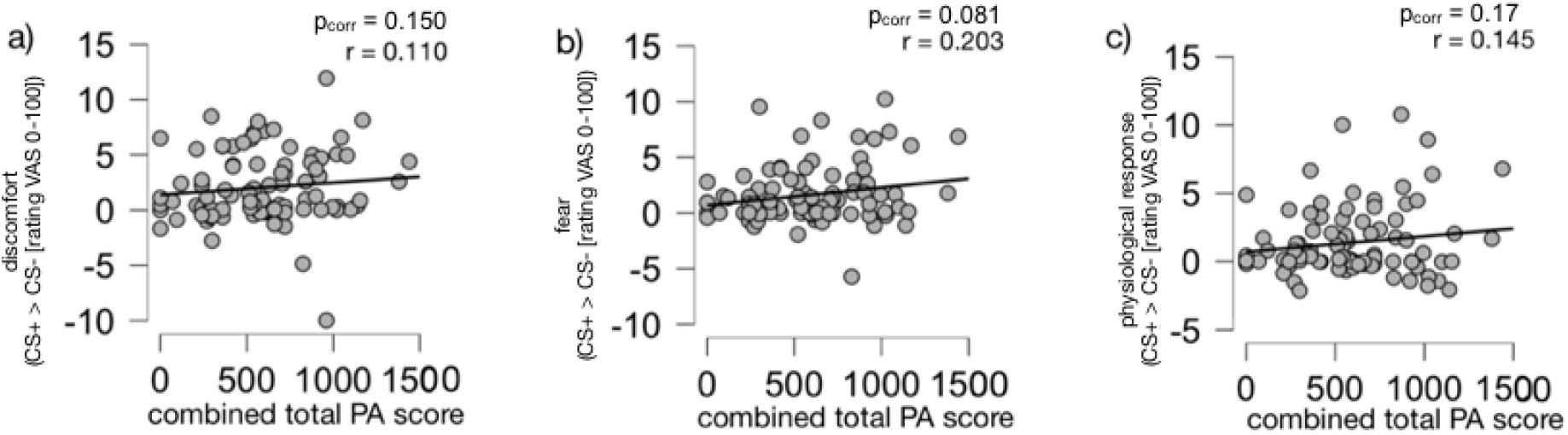
No significant correlations between the differential CS-rating (i.e., (post CS+ -pre CS+) -(post CS- - pre CS-)) of discomfort (a), fear (b) and physiological response (c) (y-axis) and the combined total physical activity score (x-axis) on day 1.

**Figure 5.**
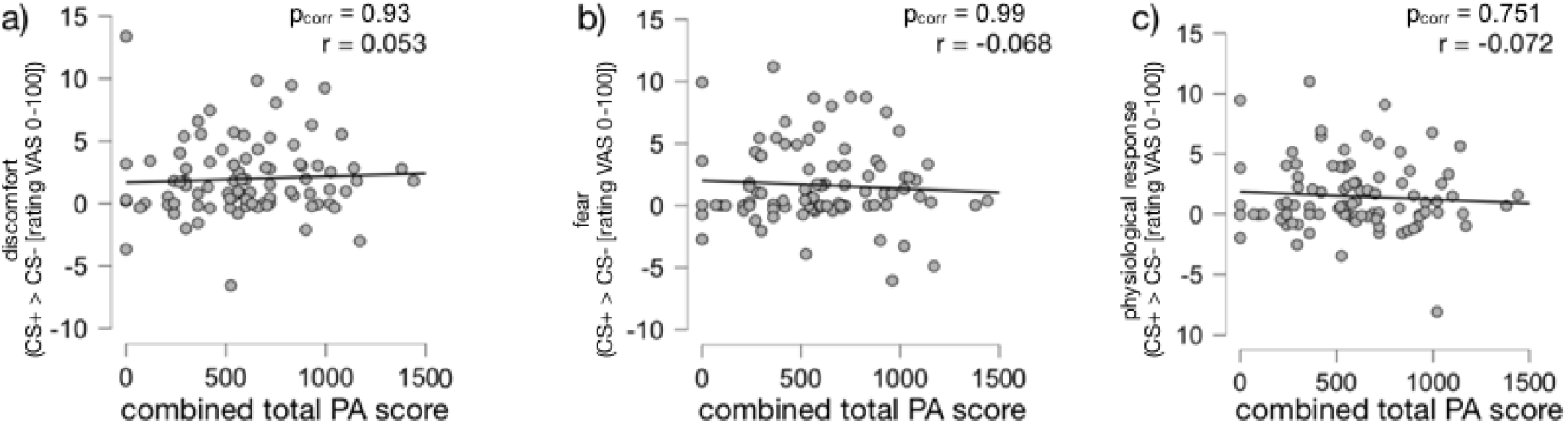
Correlations between the baseline-corrected differences of discomfort (a), fear (b) and physiological response (c) (y-axis) and the combined total physical activity score (x-axis) on day 2.

To quantify the evidence for the absence of the correlation, we calculated a Bayesian correlation analysis, which provided strong evidence for the Null-hypothesis for fear (BF_01_ = 11.850) and physiological response (BF_01_ = 12.124) on day 2, moderate evidence for the Null-hypothesis for discomfort (BF_01_ = 4.917) on day 2 and anecdotal evidence for discomfort (BF_01_ = 2.657) and physiological response (BF_01_ = 1.649) on day 1 (see Table 5). Interestingly, Bayesian correlation analysis provided no evidence for the Null-hypothesis for an association between fear ratings and PA (BF_01_ = 0.627) on day 1 (see Table 5).

**Table 5.**
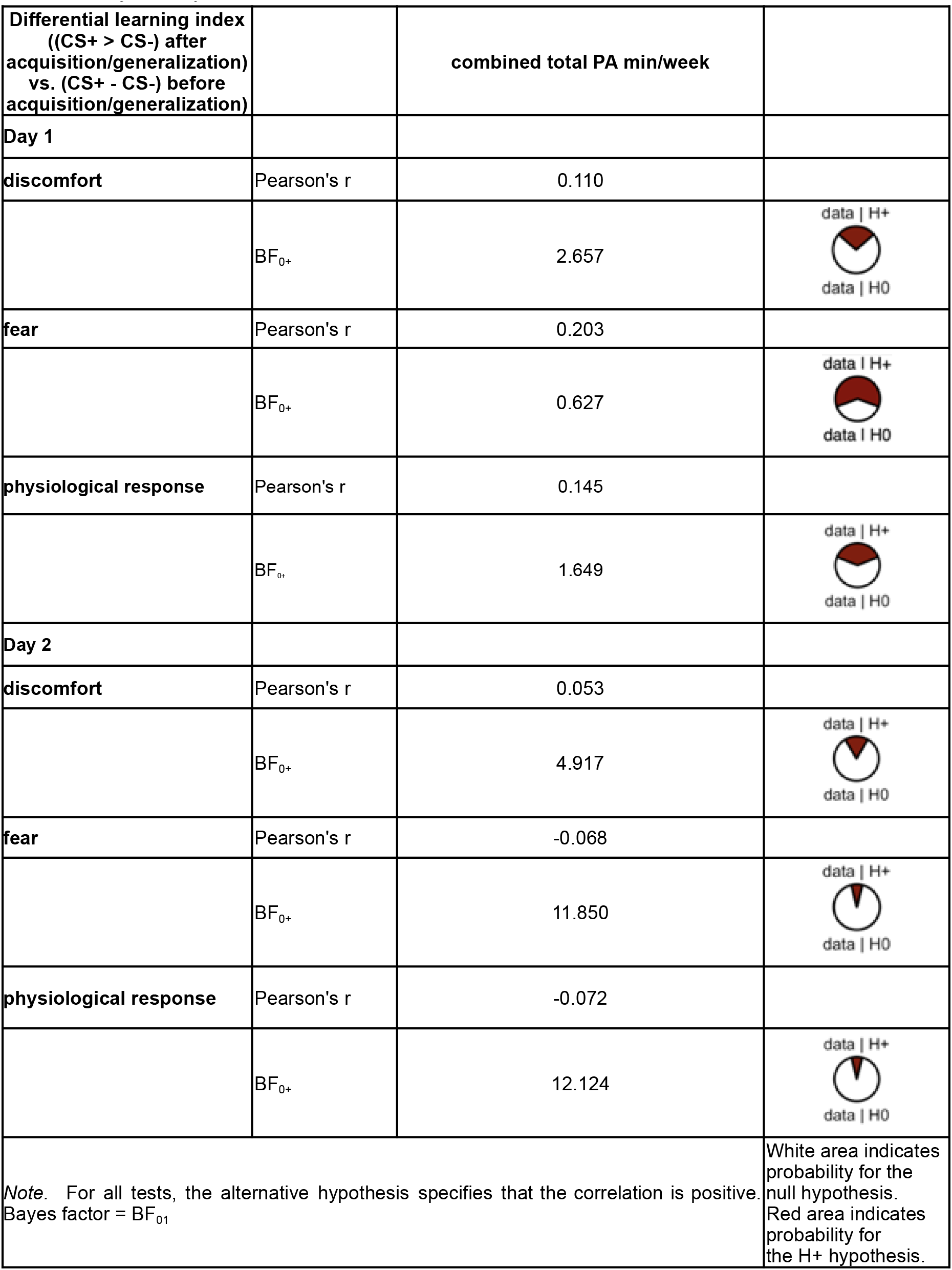
Bayesian pearson correlations with baseline-corrected differences.

| Differential learning index<br>((CS+ > CS-) after<br>acquisition/generalization)<br>vs. (CS+ - CS-) before<br>acquisition/generalization) |  | combined total PA min/week |  |
| --- | --- | --- | --- |
| **Day 1** |  |  |  |
| **discomfort** | Pearson's r | 0.110 |  |
| | $BF_{0+}$ | 2.657 | data \| H+<br>data \| H0 |
| **fear** | Pearson's r | 0.203 |  |
| | $BF_{0+}$ | 0.627 | data \| H+<br>data \| H0 |
| **physiological response** | Pearson's r | 0.145 |  |
| | $BF_{0+}$ | 1.649 | data \| H+<br>data \| H0 |
| **Day 2** |  |  |  |
| **discomfort** | Pearson's r | 0.053 |  |
| | $BF_{0+}$ | 4.917 | data \| H+<br>data \| H0 |
| **fear** | Pearson's r | -0.068 |  |
| | $BF_{0+}$ | 11.850 | data \| H+<br>data \| H0 |
| **physiological response** | Pearson's r | -0.072 |  |
| | $BF_{0+}$ | 12.124 | data \| H+<br>data \| H0 |
| **Note.** For all tests, the alternative hypothesis specifies that the correlation is positive.<br>Bayes factor = $BF_{01}$ | | | White area indicates probability for the null hypothesis.<br>Red area indicates probability for the H+ hypothesis. |

#### rmANOVA

Since the pre-registered correlation between PA and CS-differentiation did not indicate an association, but the Bayesian analyses did not provide evidence for an absence of an association (between PA and fear ratings on day 1), we explored the possible covariation between PA and CS-responses within a repeated measures analysis of variance (rmANOVA). This rmANOVA revealed significant interactions between stimulus and combined total PA (stimulus * combined total PA) for subjective discomfort, F(1.000) = 5.622, p = 0.020, η_p_^2^ = 0.063, and fear, F(1.000) = 4.827, p = 0.031, η_p_^2^ = 0.055 on day 1 and discomfort, F(1.000) = 23.743, p = 0.036, η_p_^2^ = 0.052 and fear F(1.000) = 7.158, p = 0.009 on day 2 (see table 6).

**Table 6.** Repeated measures ANOVA.

| Cases | Sum of Squares | df | Mean Square | F | p | $\eta^2_p$ |
| --- | --- | --- | --- | --- | --- | --- |
| **Day 1** |  |  |  |  |  |  |
| **stimulus (discomfort) \* combined total PA min/week** | 14.955 | 1.000 | 14.955 | 5.622 | 0.020 | 0.063 |
| **stimulus (fear) \* combined total PA** | 13.918 | 1.000 | 13.918 | 4.827 | 0.031 | 0.055 |
| **stimulus (physiological response) \* combined total PA** | 4.788 | 1.000 | 4.788 | 1.995 | 0.162 | 0.023 |
| **Day 2** |  |  |  |  |  |  |
| **stimulus (discomfort) \* combined total PA** | 23.743 | 1.000 | 23.743 | 4.565 | 0.036 | 0.052 |
| **stimulus (fear) \* combined total PA** | 31.883 | 1.000 | 31.883 | 7.158 | 0.009 | 0.079 |
| **stimulus (physiological response) \* combined total PA** | 11.668 | 1.000 | 11.668 | 2.999 | 0.087 | 0.035 |
| *Note.* Type III Sum of Squares |  |  |  |  |  |  |

### Bayesian rmANOVA

To follow upon the significant covariation of PA scores with fear and discomfort models, we calculated a Bayesian rmANOVA. These analyses allow to quantify the evidence to include the covariate of PA into a model to explain the data, when comparing to a null model that includes only the main effects of task (i.e., time, stimulus, time*stimulus, subject). These analyses indicated anecdotal evidence to include the interaction between stimulus*combined total PA score to the model for fear ratings on day 1. The analysis further provided moderate evidence for the inclusion of the combined total PA score, as well as the interaction between stimulus*combined total PA score for fear on day 2. Additionally, we found anecdotal evidence for the inclusion of an stimulus*combined total PA score for the discomfort model on day 2 (see table 7).

**Table 7.** Bayesian repeated measures ANOVA. Comparison of combined total PA to null model (incl. time, stimulus, time*stimulus, subject).

| Cases | BF<sub>incl</sub> |
| --- | --- |
| **Day 1** |  |
| combined total PA (discomfort) | 0.285 |
| stimulus* combined total PA (discomfort) | 0.541 |
| combined total PA (fear) | 0.574 |
| stimulus* combined total PA (fear) | 1.265 |
| **Day 2** |  |
| combined total PA (discomfort) | 0.598 |
| stimulus* combined total PA (discomfort) | 1.491 |
| combined total PA (fear) | 2.974 |
| stimulus* combined total PA (fear) | 8.864 |

## Discussion

### Summary

Our results provided no convincing evidence for a correlation between differential observational threat learning and physical activity (as measured by the IPAQ). However, we also found no sufficient evidence supporting the null hypothesis (i.e. no correlation) for the association between differential learning and PA on day 1, when applying bayesian statistics. Instead, we found a covariation between the ratings of discomfort and physical activity, as well as between ratings of fear and physical activity on day 1 and 2 within a repeated measurement ANOVA. This was supported by bayesian statistics.

### Previous research

Our results are in contrast to the quantity of previous research, which has demonstrated that physical activity affects learning and memory processing in rodents and humans (Chang & Etnier, 2009; Chang, Labban, Gapin, & Etnier, 2012; Coles & Tomporowski, 2008; Lambourne & Tomporowski, 2010). It also stands against the finding that exercise can enhance contextual fear acquisition in rats (Baruch et al., 2004). Our results do not indicate an association between improved differential socio-emotional learning processes and PA. However, the previous literature does not only support the hypothesis that exercise enhances all types of learning, as demonstrated by Jacquart and colleagues (2017). They showed that exercise does not facilitate fear extinction in rats. Moreover, they showed that humans do not improve their anxiety-related symptoms, when adding an exercise intervention to exposure therapy of anxiety-related disorders. Their intervention consisted of a vigorous-intensity exercise program, which was applied prior to the exposure therapy sessions. Timing and intensity of exercise may be important determinants for the effects on fear conditioning and extinction (Tanner et al., 2018). In contrast, another study that employed acute bouts of exercise by Keyan and Bryant (2019), indicated an enhancement of fear extinction.

Our study did not consider exercise interventions and might rather reflect an overall movement behavior, which might explain the reduced support for the association between PA and differential learning. Nevertheless, we found a covariation between PA and ratings of fear and discomfort, which might indicate that individual differences in physical activity can explain variance in affective ratings to discriminate between danger and threat signals.

Importantly, our findings do not indicate covariation or interactions with the factor time (i.e., a difference before vs. after learning). Hence, it is not likely that the current data indicates an association between PA and actual changes in discriminatory learning during observation threat learning. The support for the covariation with the stimulus factor rather indicates an association with the general ability to discriminate between the threatening and safe stimuli.

### Limitations

The current findings should be interpreted with caution in light of potential limitations. For the first instance the current study was conducted online and individual differences in engagement with the learning-task might have been larger, when compared to lab-based studies.

Furthermore, the current study design did not consist of an intervention of PA and therefore allows statements about associative interdependencies only. It should further be noted that a reporting-bias for the self-report of PA cannot be fully excluded.

With regard to the results of the present study it should be noted that false negatives can not be ruled out, based on the relatively conservative corrections for multiple comparisons included in the current analyses.

## Data Availability

All data produced are available online at https://gin.g-node.org

https://gin.g-node.org/MadeleineMueller/Mueller_et_al_2024_Observational_avoidance

https://gin.g-node.org/Petalas/Petalas_et_al_2024/src/master/

## Notes

### Competing Interest Statement

The authors have declared no competing interest.

### Funding Statement

This project was supported by a Young Investigator Grant from the German-Israeli Foundation (no. 2528) awarded to J.H.

### Author Declarations

Ethics committee of Aerztekammer Hamburg gave ethical approval for this work

